# Diagnosis and Severity Assessment of COPD using a Novel Fast-Response Capnometer And Interpretable Machine Learning

**DOI:** 10.1101/2023.12.20.23300327

**Authors:** Leeran Talker, Cihan Dogan, Daniel Neville, Rui Hen Lim, Henry Broomfield, Gabriel Lambert, Ahmed B Selim, Thomas Brown, Laura Wiffen, Julian C Carter, Helen F Ashdown, Gail Hayward, Elango Vijaykumar, Scott T Weiss, Anoop Chauhan, Ameera X Patel

## Abstract

**Introduction:** Spirometry is the gold standard for COPD diagnosis and severity determination, but is technique-dependent, non-specific, and requires administration by a trained healthcare professional. There is a need for a fast, reliable, and precise alternative diagnostic test. This study’s aim was to use interpretable machine learning to diagnose COPD and assess severity using 75-second carbon dioxide (CO_2_) breath records captured with TidalSense’s N-Tidal^™^ capnometer.

**Methods:** For COPD diagnosis, machine learning algorithms were trained and evaluated on 294 COPD (including GOLD stages 1-4) and 705 non-COPD participants. A logistic regression model was also trained to distinguish GOLD 1 from GOLD 4 COPD with the output probability used as an index of severity.

**Results:** The best diagnostic model achieved an AUROC of 0.890, sensitivity of 0.771, specificity of 0.850 and positive predictive value of 0.834. A potential clinical use for this model is to rule in or rule out a diagnosis in patients where the model is most confident. Performance on test capnograms with probability *>*80% and *<*20% was also evaluated, yielding a PPV of 0.930 and NPV of 0.890. The severity determination model yielded an AUROC of 0.980, Sensitivity of 0.958, Specificity of 0.961 and PPV of 0.958 in distinguishing GOLD 1 from GOLD 4. Output probabilities from the severity determination model produced a correlation of 0.71 with percentage predicted FEV_1_.

**Conclusion:** The N-Tidal^™^ device could be used alongside interpretable machine learning as an accurate, point-of-care diagnostic test for COPD, particularly in primary care, as a rapid rule-in or rule-out test. N-Tidal^™^ also could be effective in monitoring disease progression, providing a possible alternative to spirometry for disease monitoring.

**Trial registration:** Please see NCT03615365, NCT02814253, NCT04504838, NCT03356288 and NCT04939558.

## Introduction

According to the World Health Organization in 2020, Chronic Obstructive Pulmonary Disease (COPD) was the third leading cause of mortality, accounting for 6% of global deaths [1]. It is estimated that the prevalence of COPD is growing, with the number of people receiving a COPD diagnosis in the UK alone increasing by 27% in the past 10 years [2]. This trend is only set to increase over the next decade [3]. Currently a cure for COPD does not exist [4]. Therefore, early diagnosis and treatment is key to prevent disease progression, improve quality of life, reduce exacerbations, and limit the economic burden associated with management of the disease [5, 6]. Spirometry is the current gold standard for diagnosis and COPD severity determination, but it relies heavily upon a patient’s ability to exhale forcefully. There is also a widespread shortage of trained healthcare professionals able to perform quality assured spirometry and its use remains limited in some areas due to the associated risk of an aerosol-generating cough. In addition, spirometry has been shown to be ineffective at screening for early cases [7]. It has been estimated that in the UK, only between 9.4% and 22% of those with COPD have been diagnosed [8], in part due to spirometry’s poor precision of only 63% [9]. The global shortage of adequately trained staff and protracted waiting lists for spirometry have resulted in the Lancet Commission recognizing that diagnostics must move beyond spirometry [10].

Capnography is a widely used technique in critical care and anesthesia. A previous study by Talker et. al. [11] showed that features derived from a patient’s high-resolution capnogram using the TidalSense N-Tidal^™^ device can be used to diagnose severe COPD with a high degree of accuracy, suggesting the technique could be an appealing alternative to spirometry. The COPD diagnosis element of this study built on previous analysis by adding patients with less severe COPD to the dataset, increasing the resulting model’s real-world applicability.

The objective of this study was to apply interpretable machine learning techniques to capnography data recorded by the N-Tidal^™^ device across five clinical studies and evaluate the performance of the diagnostic and severity classifiers. The primary aim was to construct a classifier that could distinguish capnograms of patients with varying severities of COPD (GOLD stage 1-4) from those without COPD. A secondary aim was to develop an alternative severity index to percentage predicted FEV_1_ which could be used as an aid by clinicians to quantify COPD severity. The potential advantages of using the N-Tidal^™^ device in the context of COPD diagnosis include: the capture of CO2 data from tidal breathing for improved ease of use and absence of aerosol-generating cough; the speed of administration of the test (under five minutes); and an automated diagnostic output not reliant on specialist training [11]. These advantages could extend to using the device for severity determination and could be helpful clinically in monitoring disease progression.

## Methods

### Studies

The capnography data used for this analysis were collected from five different longitudinal observational studies, namely CBRS, CBRS2, GBRS, ABRS and CARES. A summary of each of these studies containing their objectives, participant information and inclusion/exclusion criteria can be found in the supplementary material. These studies and therefore the dataset used in this analysis included patients with COPD (GOLD stage 1-4), asthma, heart failure, pneumonia, breathing pattern disorder, motor neuron disease, sleep apnea, bronchiectasis, pulmonary fibrosis, tracheobronchomalacia, anemia, lung cancer, long COVID-19, general upper airway obstruction, extrinsic allergic alveolitis, as well as healthy participants. The heterogeneity of diseases present in the dataset ensured the resultant model’s generalizability to a real-world diagnostic scenario, where patients would be expected to present with a variety of cardiorespiratory and other conditions which have similar initial symptoms.

In patients with COPD, diagnoses were made according to NICE guidelines. COPD severity (GOLD stage 1-4) was determined from the percentage predicted FEV_1_ of the subset of COPD patients where spirometry was available. Diagnostic criteria used for other conditions, including asthma, are in the supplementary material. Potential participants were identified in outpatient clinics, inpatient wards, primary care and secondary care clinics according to each study’s protocol before undergoing a screening process with the study team to assess their suitability.

Alongside the five studies noted above, capnography data was collected from 72 volunteers without any respiratory disease between December 2015 and May 2022. These healthy volunteers provided written informed consent and were screened by a doctor to ensure they did not have any confounding cardiorespiratory disease or other comorbidities. All subjects across the five studies gave informed consent, and their data was handled according to all relevant data protection legislation, including the EU/UK General Data Protection Regulation (GDPR).

Ethical approval was obtained from the South Central Berkshire Research Ethics Committee (REC) for GBRS and ABRS, the Yorkshire and the Humber REC for CBRS and CARES, and the West Midlands Solihull REC for CBRS2.

### Procedure

In all studies, capnography data was collected using the N-Tidal^™^ device, a CE-marked medical device regulated in the UK and EU. N-Tidal^™^ has been designed to take accurate, reliable recordings of respired pressure of CO2 (pCO2) directly from the mouth. It is unique in its ability to accurately measure pCO2 from ambient/background CO2 to hypercapnic levels in exhaled breath through unforced tidal breathing with a fast response time, meaning that quick changes in the geometry of the pCO2 waveform are captured. CO2 was sampled at 10kHz and reported at 50Hz providing a level of resolution not possible with alternative capnometers. This resolution was critical to the machine learning models’ ability to use subtle geometric features of the waveform to distinguish capnograms [12].

Study participants were given an N-Tidal^™^ device to take home after completing training on the correct operation and storage of the device. Capnography data was serially collected twice daily for varying lengths of time ranging from 2 weeks up to 12 months according to each study’s protocol. Patients performed normal tidal breathing through the device for 75 seconds through a mouthpiece in an effort-independent process. A single episode of use (breath recording) produced a single capnogram, with each respiratory cycle (inspiration and expiration) forming a single waveform. In addition to capnometry data, the following data was also collected from the majority of participants on all five studies: basic demographics, spirometry, smoking history, comorbidities and medications. Some of this information was used in conjunction with capnography data in subsequent analysis. Other clinical and questionnaire data varied across studies (see supplementary material). Personal data was pseudonymized in accordance with GDPR.

### Feature engineering

Raw pCO_2_ data collected in each breath recording was first denoised then separated into breaths, each of which was then segmented into its constituent phases. At this point, breaths which were anomalous and could not be processed were excluded from subsequent processing and analysis using a set of proprietary rules built into the N-Tidal cloud platform.

To generate features for machine learning classification, two categories of information were captured: geometric characteristics of the waveform associated with each breath (referred to as ‘per breath features’); and features of the whole capnogram, such as respiratory rate or maximum end-tidal CO_2_ (ETCO2), referred to as ‘whole capnogram features’.

Any breaths where the full feature set could not be calculated were also automatically excluded from analysis, and further checks were carried out manually to ensure that all condensation-compromised breaths had been excluded by automated methods.

### Machine learning

Following pre-processing and waveform parameterization, a reduced number of capnograms were randomly selected for each participant in the dataset such that the COPD and non-COPD cohorts had the same number of capnograms. Then, each machine learning model was trained and evaluated using a nested cross-validation scheme (see supplementary material for more information).

### COPD diagnosis

N-Tidal capnographic features were used to train three different machine learning algorithms: logistic regression (LR); extreme gradient boosted trees (XGBoost); and a support vector machine (SVM). LR and XGBoost were chosen as they are very different in their approach to training yet similarly interpretable, while SVM was chosen as the non-linear shallow-learning algorithm which often maximizes supervised performance.

Model performance was delineated into performance on each of the most commonly presenting diseases in the non-COPD class and each severity in the COPD class. The impact of the most prevalent comorbidities on model performance was assessed for participants with comorbidities in addition to COPD, to investigate the model’s analogy to a real-world use case where testing on patients with COPD and additional comorbidities is likely to be common. The most significant features driving model learning were extracted to understand which areas of the capnogram waveform were most predictive of COPD.

### Severity determination

A proposed method for severity determination was to train a machine learning classifier to distinguish between participants with GOLD 1 and GOLD 4 COPD and use the probability of having GOLD 4 outputted by this model as an indication of COPD severity. This ‘severity index’ would indicate where on the spectrum between mild and very severe COPD a patient falls. When testing on unseen patients of all severities, the output probability distribution for GOLD 2 and GOLD 3 patients would be expected to fall in between the distributions for GOLD 1 and GOLD 4, and for a patient’s probability output (severity index) to steadily increase as their disease progresses.

Given the small number of patients available for this analysis and the predominance of comorbidities especially among GOLD 1 patients (25/47 patients), this analysis was conducted after removing comorbid patients from the dataset. This was so the machine learning model would be able to more effectively infer the underlying signal of COPD severity without the potential confounder of comorbidities. Following model training, the distribution of each COPD severity in the model’s probability output was investigated, and random waveforms from adjoining severity classes were inspected to explore the probability output’s efficacy as a tool for severity determination. To investigate the relationship between traditional spirometric methods and the proposed model probability severity indication, each patient’s severity model probability output was plotted against their paired percentage predicted FEV_1_ value and a correlation was calculated.

## Results

Between 6 December 2015 and 8 December 2022, 115,053 capnograms were collected from 1114 patients. On average, each patient collected 103 capnograms over 57 days. Demographic data for the preprocessed and class-balanced machine learning dataset were collated (Table 1).

**Table 1.** Demographic information from the five studies and the separate healthy volunteer cohort. Categorical data are given as a number with its percentage of the total (n (%)). Continuous data given as (median (Q1-Q3)). For the COPD classification dataset, smoking history and pack years were absent for 82 and 281 participants respectively. For the severity determination dataset, smoking history and pack years were absent for 2 and 16 participants respectively.

|  | Diagnosis |  |  | Severity |  |  |
| --- | --- | --- | --- | --- | --- | --- |
|  | COPD<br>(N=294) | Non-COPD<br>(N=705) | Overall<br>(N=999) | Mild COPD<br>(N=22) | Very Severe COPD<br>(N=21) | Overall<br>(N=43) |
| **Age** | 67 (60-74) | 54 (44-66) | 59 (48-69) | 68 (59-75) | 66 (61-69) | 66 (60-74) |
| **Gender (Female)** | 138 (47.1%) | 395 (62.7%) | 533 (57.8%) | 8 (36.4%) | 9 (40.9%) | 17 (39.5%) |
| **BMI (kg/m<sup>2</sup>)** | 26.4 (23.0-32.3) | 27.6 (23.7-32.8) | 27.2 (23.1-32.6) | 25.3 (23.0-30.5) | 25.4 (22.1-29.5) | 25.4 (22.2-29.6) |
| **Smoking History:** |  |  |  |  |  |  |
| Current smoker | 61 (21.1%) | 51 (8.1%) | 112 (12.2%) | 7 (31.8%) | 1 (5.3%) | 8 (19.5%) |
| Ex-smoker | 207 (71.6%) | 248 (39.4%) | 455 (49.6%) | 12 (54.5%) | 18 (94.7%) | 30 (71.1%) |
| Never smoked | 21 (7.2%) | 329 (52.4%) | 350 (38.2%) | 3 (13.6%) | 0 (0.0%) | 3 (7.3%) |
| **Pack Years (all)** | 30.8 (15.0-42.3) | 0.0 (0.0-6.1) | 2.1 (0.0-19.4) | 25.0 (5.0-37.5) | 54.5 (37.5-74.8) | 35.0 (17.8-51.8) |
| Current smoker | 33.8 (28.9-39.9) | 15.8 (8.8-30.0) | 28.5 (12.0-38.1) | 33.0 (20.6-45.2) | 45.0 (45.0-45.0) | 39.0 (26.8-48.1) |
| Ex-smoker | 32.3 (18.8-45.0) | 6.7 (2.0-15.0) | 15.0 (4.1-31.4) | 30.0 (17.8-39.4) | 63.0 (35.0-75.0) | 36.3 (25.0-64.8) |

### COPD diagnosis

All performance statistics were obtained using a decision boundary of 0.5 on the models’ probability outputs. In classifying COPD vs non-COPD participants, the support vector machine (SVM) marginally showed the best performance, with accuracy, AUROC, sensitivity, specificity, NPV and PPV at 0.811, 0.890, 0.771, 0.850, 0.792 and 0.834 respectively (Table 2). Model performance was consistent between iterations (Table 2); all three algorithms had a standard deviation of less than 0.03 in accuracy over the five folds. These metrics were produced from the aggregated predictions on all five unseen outer-loop test sets, while the standard deviation describes the variation of model performance between the five outer-loop test sets and gives an indication of model generalizability. As performance for all models was extremely similar, further analysis is only presented for the LR model, as it is the most explainable. Table 3 outlines LR model performance of the most commonly presenting primary diseases in the non-COPD class and each GOLD severity in the COPD class. Comorbid COPD patients have been included in the performance statistics for their respective COPD severity. Notably high performance was seen in classifying healthy, asthma, breathing pattern disorder, and GOLD 2-4 COPD participants, while lower performance was seen in the heart failure and GOLD 1 COPD cohorts.

**Table 2.** Aggregated machine learning model performance on all 5 unseen outer-loop test sets, for each of the three models built: logistic regression (LR), extreme gradient boosted trees (XGBoost), and support vector machine (SVM) with an RBF kernel. The highest performance (across all models), for each of the metrics reported is highlighted in bold.

|  | Accuracy | AUROC | Sensitivity | Specificity | Negative Predictive Value (NPV) | Positive Predictive Value (PPV) |
| --- | --- | --- | --- | --- | --- | --- |
| **LR** | 0.809 ± 0.028 | 0.884 ± 0.023 | **0.788 ± 0.055** | 0.829 ± 0.017 | **0.800 ± 0.041** | 0.818 ± 0.017 |
| **XGBoost** | 0.806 ± 0.018 | **0.891 ± 0.015** | 0.771 ± 0.034 | 0.840 ± 0.012 | 0.790 ± 0.023 | 0.825 ± 0.014 |
| **SVM** | **0.811 ± 0.022** | 0.890 ± 0.020 | 0.771 ± 0.038 | **0.850 ± 0.012** | 0.792 ± 0.026 | **0.834 ± 0.015** |

**Table 3.** Accuracy of COPD/non-COPD diagnosis for most commonly presenting disease groups and all COPD severities, over all five outer-loop test sets.

|  | Number of patients | Number of capnograms | Diagnostic accuracy |
| --- | --- | --- | --- |
| **Asthma** | 257 | 1538 | 0.839 ± 0.027 |
| **Healthy** | 125 | 607 | 0.923 ± 0.056 |
| **Heart Failure** | 65 | 389 | 0.661 ± 0.055 |
| **Breathing Pattern Disorder** | 10 | 60 | 0.950 ± 0.075 |
| **GOLD 1 COPD** | 47 | 632 | 0.606 ± 0.152 |
| **GOLD 2 COPD** | 107 | 1451 | 0.815 ± 0.054 |
| **GOLD 3 COPD** | 53 | 725 | 0.898 ± 0.099 |
| **GOLD 4 COPD** | 24 | 325 | 0.985 ± 0.019 |
| **COPD (no severity label)** | 55 | 720 | 0.692 ± 0.067 |
| **Emphysema** | 8 | 112 | 0.804 ± 0.263 |

Figure 4 shows the diagnostic performance for the capnograms with a greater than 0.8 and less than 0.2 probability of COPD. These are capnograms which have been classified with high confidence by the model.

**Figure 1.**
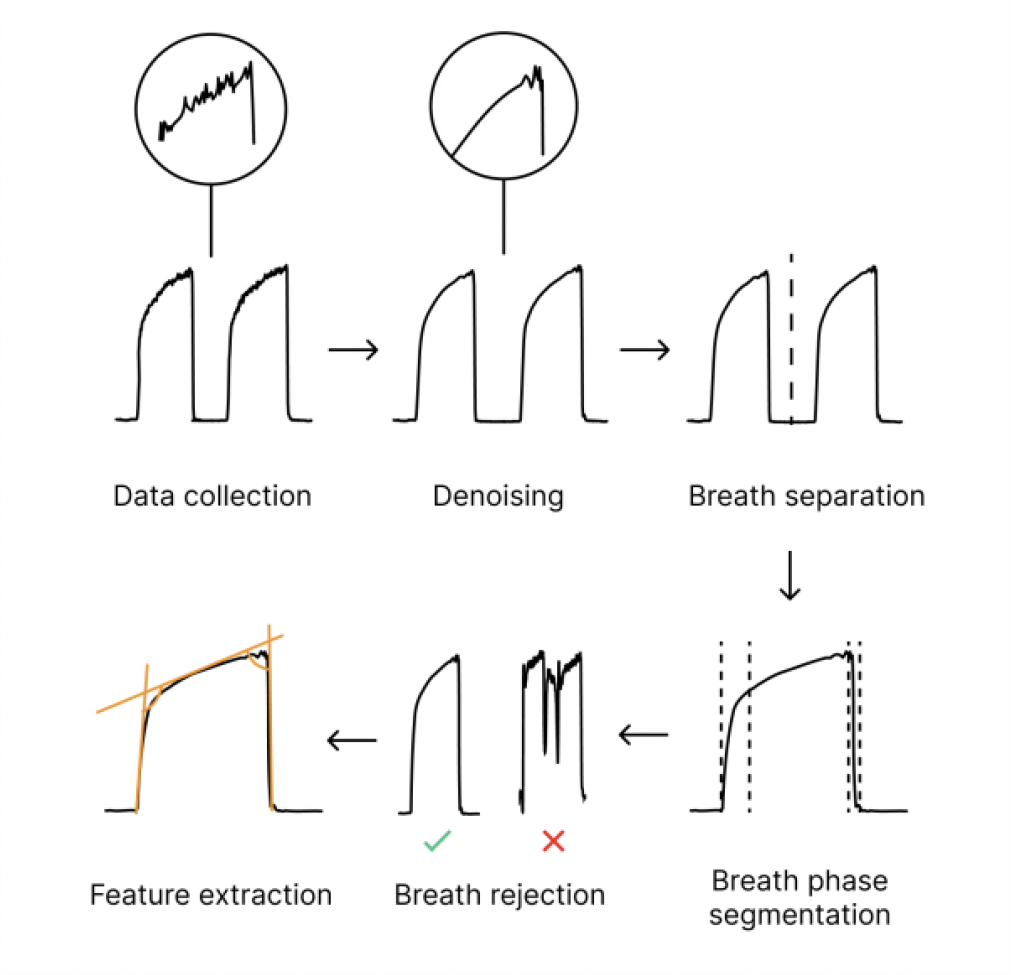
High-level overview of the data processing pipeline applied to the fast-response CO_2_ data collected through the N-Tidal^™^ device.

**Figure 2.**
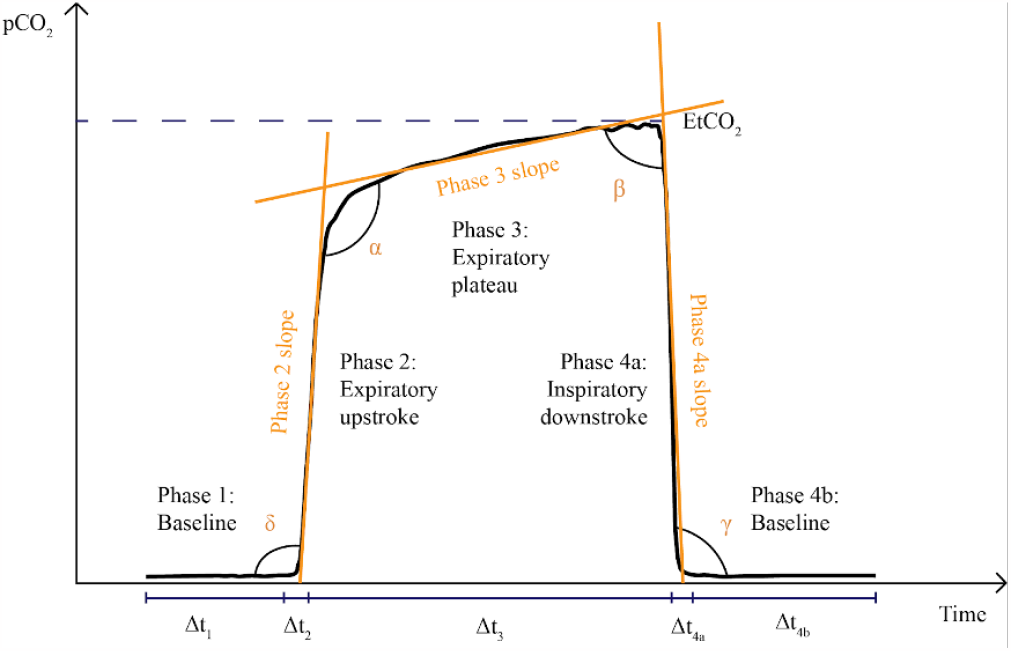
Illustration of a capnogram waveform and its phases and angles. Phase 1 is the inspiratory baseline, Phase 2 is the expiratory upstroke (representing the first phase of exhalation), Phase 3 is the expiratory plateau (representing the majority of exhalation), Phase 4a is the inspiratory downstroke (representing the first phase of inhalation), and Phase 4b is the inspiratory baseline. Note that the start of Phase 1 and the end of Phase 4b may technically be considered part of the same phase.

**Figure 3.**
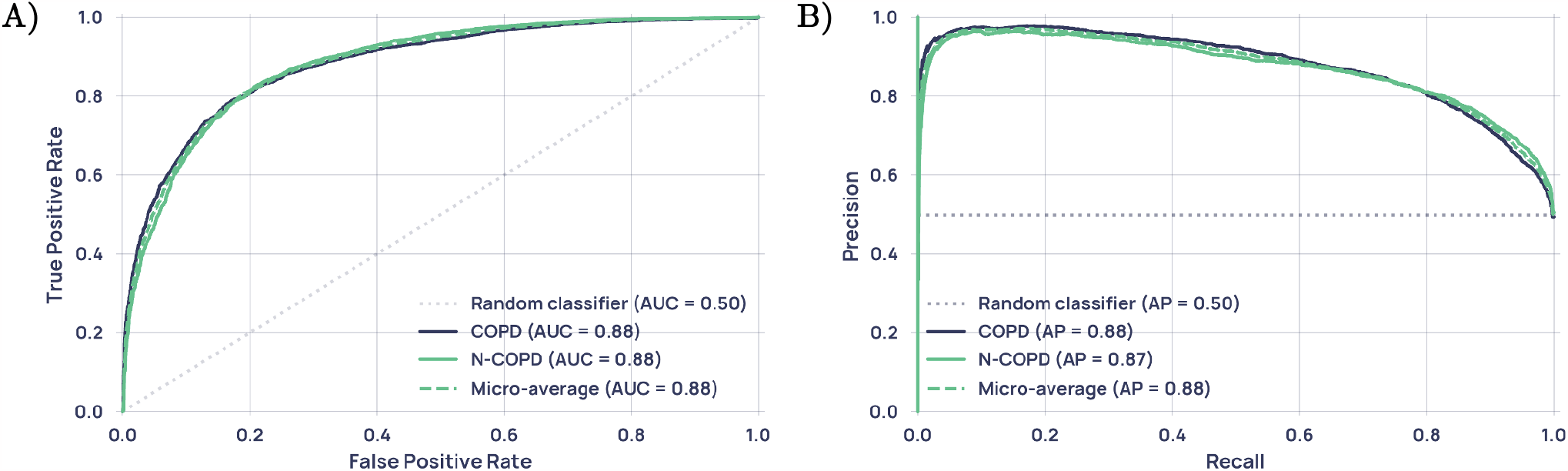
**(A)** Receiver operating characteristic (ROC) curve for the LR model, reported with results of a theoretical ‘random’ classifier with no predictive power. **(B)** Precision-Recall Curve for the LR model, reported with the results of a theoretical ‘random classifier’ and the average precision (AP).

**Figure 4.**
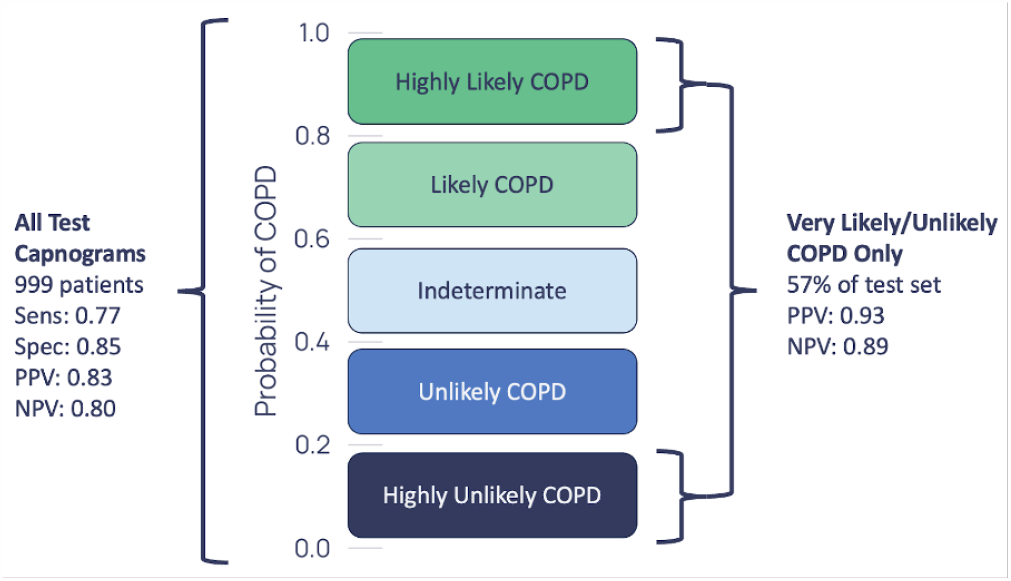
Diagnostic performance on the full aggregated test set and the highly confident regions only.

To investigate how comorbidities impacted the accuracy of COPD classification, COPD patients were grouped according to the most prevalent comorbidities and the performance was assessed for each of these groups (Table 4).

**Table 4.** Diagnostic accuracy and standard deviation across test-sets for the most prevalent COPD comorbidities. Comorbid COPD patients who had Bronchiectasis/HF/Long COVID/Pneumonia and other lung conditions were placed in the COPD and Other(s) category.

|  | Num. patients | Accuracy |
| --- | --- | --- |
| COPD and Asthma | 51 | 0.771 ± 0.095 |
| COPD, Asthma and Other(s) | 29 | 0.663 ± 0.127 |
| COPD and Bronchiectasis | 10 | 0.679 ± 0.464 |
| COPD and Long COVID | 8 | 0.830 ± 0.208 |
| COPD and Heart Failure | 5 | 0.971 ± 0.025 |
| COPD and Pneumonia | 3 | 0.865 ± 0.393 |
| COPD and Other(s) | 13 | 0.626 ± 0.416 |
| All Comorbid COPD | 119 | 0.736 ± 0.092 |

An evaluation of the predictive model was conducted to identify which capnogram features best distinguished patients with and without COPD. The relative feature importances for driving learning in the LR model were determined and the region of the capnogram waveform from which the features were extracted was used to construct the importance map for non-COPD and COPD waveforms in Figure 5. Features associated with the alpha region of the capnogram waveform as well as phase 4a (the inspiratory downstroke) were found to be the most important drivers of learning.

**Figure 5.**
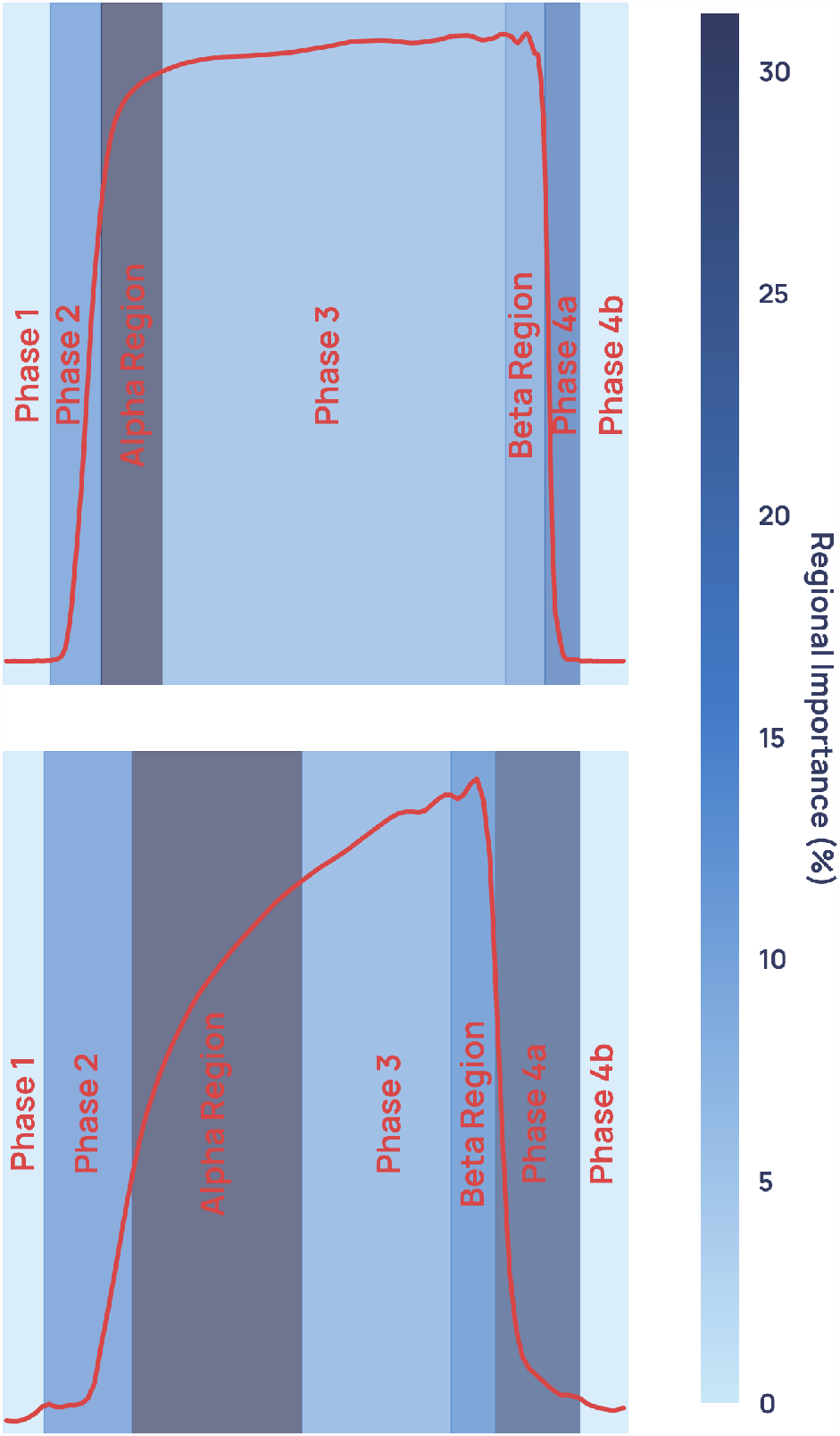
Average weighted feature importance for COPD diagnosis by capnogram waveform region, where weighted features were calculated as the magnitude of the product of the standardized feature value and the feature importance. **(A)** shows an example for a non-COPD waveform, and **(B)** shows an example for a COPD waveform.

### Severity determination

Classification was performed using only logistic regression in this analysis due to its simplicity, explainability and since logistic regression was shown in the COPD diagnostic task to be as accurate as non-linear models.

All performance statistics were obtained using a decision boundary of 0.5 on the models’ probability outputs. The classification model trained to distinguishing GOLD 1 from GOLD 4 participants achieved an accuracy of 0.959 ± 0·039, an AUROC of 0.980 ± 0·013, a sensitivity of 0.958 ± 0·051, a specificity of 0.961 ± 0·045, NPV of 0.961 ± 0·047, and PPV of 0.958 ± 0·039 in, where the standard deviation describes the variability of performance on the unseen outer-loop test sets across the five folds.

Manual inspection of the average capnogram waveform of each GOLD severity (Figure 6) demonstrates that greatest visual difference between severities is seen in the transition between the expiratory upstroke and expiratory plateau, along with marked differences in the beta region and in the phase 2/4a regions. In addition, it can be seen that the difference in the average waveform between the GOLD 1 and GOLD 2 waveforms is slight compared to the difference between GOLD 2 and GOLD 3.

**Figure 6.**
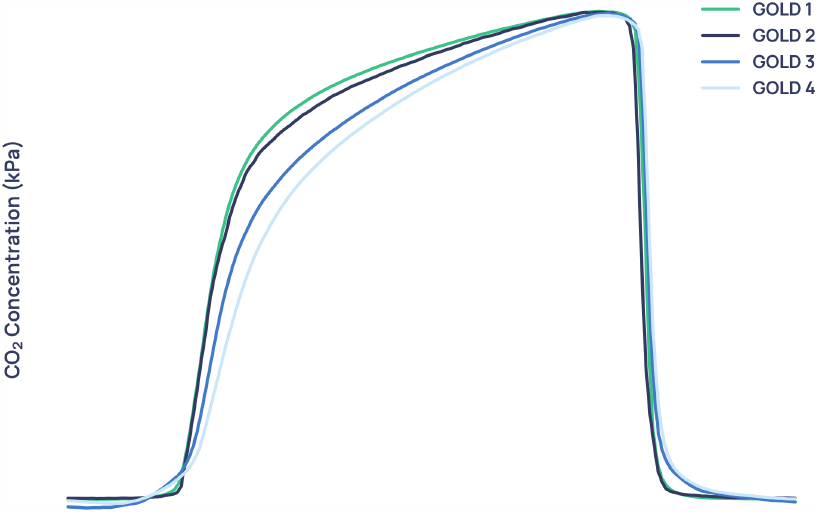
Capnogram waveforms averaged across all patients of each severity in the training dataset and normalized to equal width and height.

The model which gave the median accuracy on the unseen outer-loop test set out of the five folds was chosen for further analysis. It was used to output prediction probability for all GOLD 2 and GOLD 3 patients’ capnograms, the remainder of GOLD 1 and GOLD 4 capnograms belonging to patients who were unseen for this particular model, as well as all comorbid COPD patients whose severity could be inferred from percentage predicted FEV_1_. Figures 7, 8 and 9 are the result of this process. Figure 7 shows the average waveforms of two GOLD 1 and two GOLD 2 capnograms.

**Figure 7.**
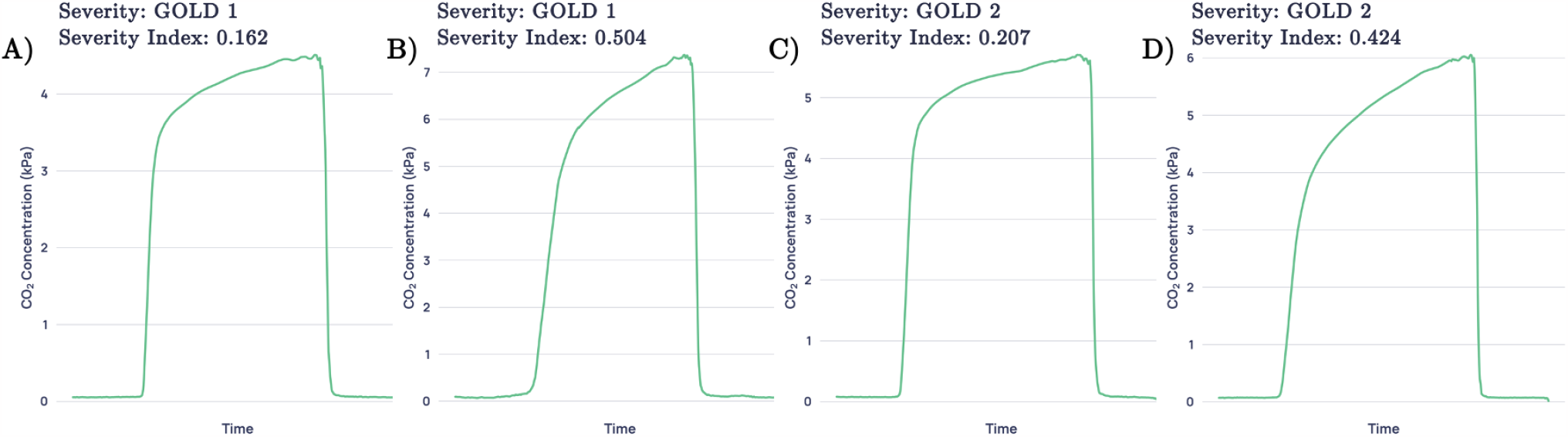
Average waveforms, where **(A)** and **(B)** are two GOLD 1 examples with the corresponding confidences and **(C)** and **(D)** show two GOLD 2 examples with the corresponding confidences. **(A)** and **(C)** are examples with low prediction confidence for their severity and **(B)** and **(D)** are examples with high prediction confidence for their severity.

**Figure 8.**
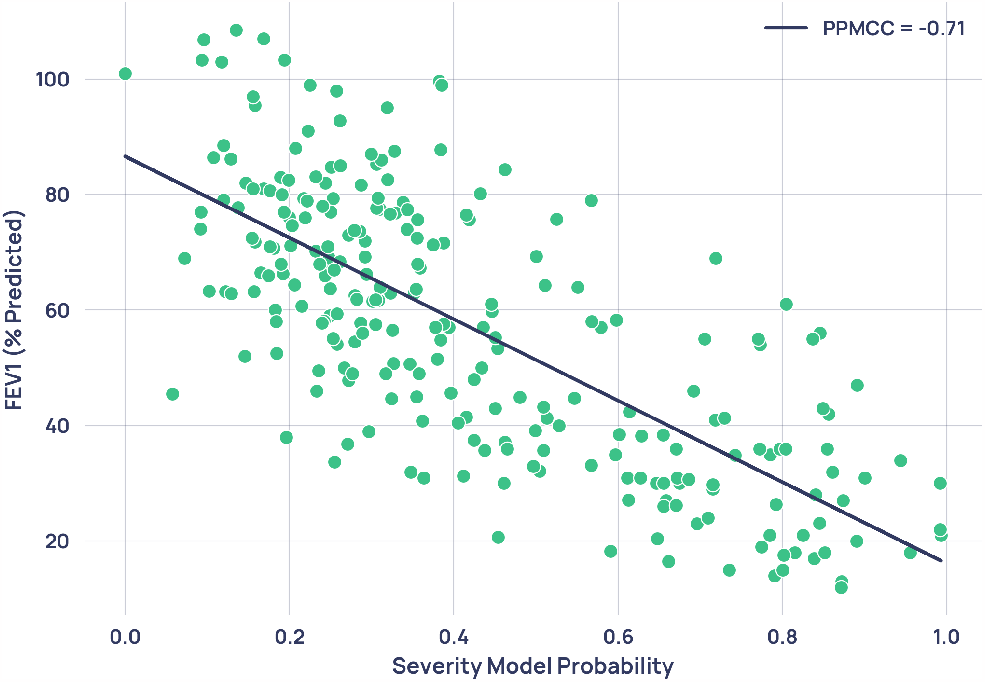
Scatterplot of severity model output probability against percentage predicted FEV_1_ from paired spirometry data. Each point represents a single paired capnogram. The correlation coefficient was -0.71.

**Figure 9.**
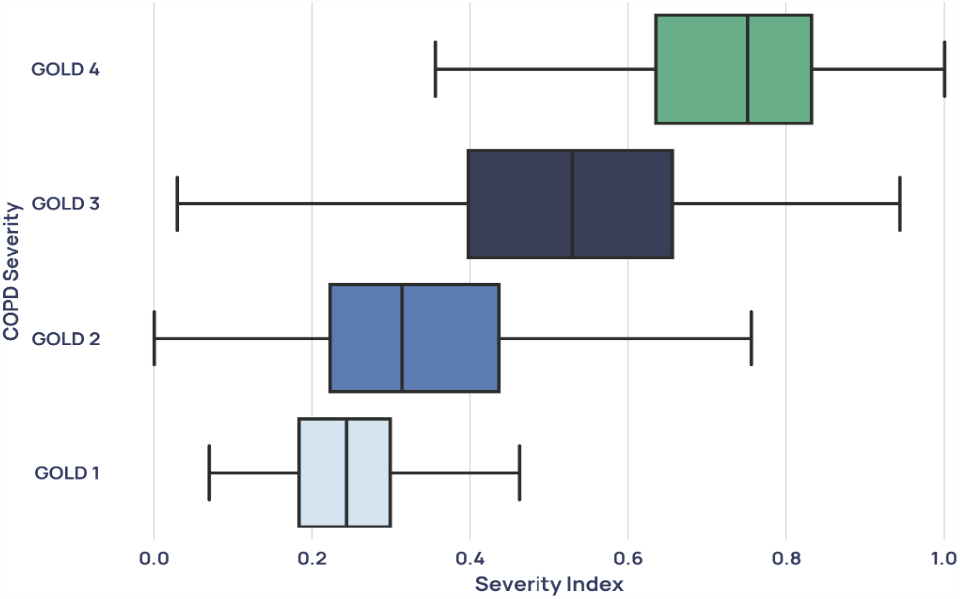
Boxplot showing the distribution of each GOLD stage in the severity model’s probability output.

For each severity, one capnogram with probability below the lower quartile was chosen and one capnogram with probability above the upper quartile was randomly chosen. To understand the relationship between the standard spirometric indication for COPD severity and the severity index developed in this paper, the percentage predicted FEV_1_ for each participant was correlated with the severity model output probability for all 224 patients with paired spirometry (spirometry taken on the same day as a capnogram), unless the paired capnogram was used for training. This data was plotted in Figure 8, where the linear relationship gave a Pearson’s product moment correlation coefficient (PPMCC) of -0.71.

Figure 9 shows the distribution of each GOLD severity in the severity model’s probability output, including capnograms which were unpaired with spirometry. These unpaired capnograms were designated the severity given by the patient’s median percentage predicted FEV_1_ measurement.

## Discussion

### Model robustness

For the diagnostic task, the best performing model over the five unseen outer-loop test sets was the support vector machine (SVM), with a class-balanced AUROC of 0.811 ± 0·022 and positive predictive value (PPV) of 0·834 ± 0·015. The robustness of the diagnostic models was demonstrated through the similar performances of all three algorithms, showing a strong signal in the data itself; the small difference between training set and testing set diagnostic accuracy (0.819 vs 0.809 for logistic regression, for example); and the low variability of performance across different test sets, showing the model’s generalizability to a true clinical scenario. See supplementary material for more details.

### Interpretability and waveform features driving learning

The identified features (Figure 5) contributing most to the logistic regression model’s decision were from the *α* angle region, which characterizes the rate at which gas from the upper airways (CO_2_-poor) gives way to mixed alveolar gas from the lower airways (CO_2_-rich). A larger *α* angle corresponds to greater airway resistance, likely due to an obstructed bronchospastic airway or alveolar damage associated with emphysema.

For severity determination, the high correlation of 0.71 between the model’s probability output and the current standard for spirometric severity determination, percentage predicted FEV_1_, indicates the physiological plausibility of the severity index in describing the progression of COPD pathology.

Figure 6 shows the average waveforms for each COPD severity. We observe that the more severe forms of COPD exhibit a more extreme “shark-fin” like shape, characterized by a large *α* angle and more slanted expiratory plateau. The shark-fin type waveform is known to arise from differences in time constants of gas movement from different alveoli to the sensor, as a result of differing compliance and resistance of alveoli. The changes in compliance and resistance arise as a result of the airway remodeling that occurs in COPD. This changes the rate of transition of gas from alveoli to anatomical dead space (i.e. the expiratory upstroke). Notably, the average waveforms of GOLD 1 and GOLD 2 waveforms are extremely similar, with greater distinction between GOLD 2/GOLD 3, and GOLD 3/GOLD 4 waveforms. This similarity is reflected in the probability distributions of the mild and moderate severity groups (Figure 9), which overlap more than severe and very severe patients do. One possible cause for the conflation of the two severities in our dataset is that GOLD 1 COPD in particular is heavily underdiagnosed using the current standard [7], and rates of misdiagnosis are also high. Therefore, misdiagnosed GOLD 1 patients in the dataset who actually had a different obstructive lung condition could have distorted the resultant average waveform, and the patients who do truly have GOLD 1 COPD were likely to be on the more severe end of their severity.

There is a subset of mild COPD patients who exhibit a “shark-fin” like waveform which resembles more severe COPD - such as example B in Figure 7. It is becoming more widely accepted that percentage predicted FEV_1_ alone does not effectively represent the functional impairment experienced by individual patients [13], a fact which catalyzed the introduction of GOLD’s ABCD and ABE systems of severity classification. It is therefore possible that high-resolution capnography is able to identify a severity signal in COPD which correlates more closely to clinical outcomes. This hypothesis would explain why GOLD 1 waveforms with “shark-fin” shapes, such as example B in Figure 7, were given a capnographic severity index which was higher than most GOLD 2 and many GOLD 3 participants. To verify this hypothesis, further analysis would be required to examine the clinical outcomes of patients who are given a higher severity index than their GOLD status would suggest.

### Diagnostic performance and model bias

The classification performance for the most commonly presenting disease groups and all COPD severities on the diagnostic task can be found in Table 3 and the performance of various categories of comorbid COPD patients can be found in Table 4. A previous study by Talker et. al [11] found that the highest performing cohorts had capnograms at the extremities of a square-shaped healthy waveform with smaller *α* angle and flatter expiratory plateau, or highly shark-fin-like waveforms with larger *α* angle and steeper expiratory plateau for COPD. These cohorts perform similarly well in this analysis, with the Healthy, GOLD 3 and GOLD 4 groups all scoring at or above approximately 90% in classification accuracy. While GOLD 2 COPD also performed well (0.815), the GOLD 1 COPD cohort performed significantly worse (0.616). Mean pack years for the misclassified and correctly classified GOLD 1 patients were 14.4 and 27.3 years respectively (Mann-Whitney U=16152, P = 1 × 10-13, two-tailed) which could suggest that the rate of mislabeling in the misclassified cohort was much higher.

Other factors which may have contributed to this result include the high prevalence of confounding comorbidities in this group (25/47 GOLD 1 patients) and that the COPD signal is weaker in GOLD 1 patients and therefore more easily confused with other conditions. A lower performance than expected was also seen in the smaller COPD group that did not have spirometry from which severity could be inferred. Several factors may have contributed to this poorer performance. First, this cohort had the highest prevalence of comorbidities among all of the disease/severity groups (31/55 patients), which could have confounded classification. Second, 95% of these patients were recruited as part of the CARES study, which was designed to specifically target milder COPD patients and therefore could have led to decreased performance. The mean pack year value of 28.9 in this group, less than the mean pack years for Moderate COPD (30.1), corroborates this hypothesis.

Notably, the classification accuracies of the asthma group (0.839) and the COPD & asthma comorbidity group (0.771) were both high, suggesting the model can effectively identify the COPD component in asthma-COPD overlap and distinguish it from pure asthma. While performance in classifying comorbid patients with COPD & Heart Failure was extremely high (0.971), performance in classifying pure Heart Failure as non-COPD was significantly lower (0.661). It is possible that a portion of cases labeled as pure Heart Failure were the result of undiagnosed COPD, which can cause right-sided heart failure by inducing pulmonary hypertension through low blood oxygen levels [14]. The mean pack years of 16.2 in the pure Heart Failure group, significantly higher than the mean of 6.3 in the non-COPD cohort overall, corroborates this.

Reassuringly, there was no significant association between prediction accuracy and demographic features (including age, birth sex, and BMI) through non-parametric statistical testing (see supplementary material), indicating that there is no systematic bias in model performance with respect to demographic data collected on the studies.

### Potential clinical applications

When presented with the capnogram of a new patient, the diagnostic model presented above outputs a probability of COPD. Rather than relying on individual clinicians to interpret what is a novel, AI-derived diagnostic output, it could be clinically useful to create categories of model confidence (Figure 4). For instance, those above a probability threshold of COPD of 0.8 could be said to be in a ‘highly likely’ category since the PPV associated with this group is 0.93 - patients within this group could potentially be immediately diagnosed with COPD and appropriate treatment initiated. Conversely, patients with a probability of under 0.2 could be said to be in a ‘highly unlikely’ category with an NPV of 0.89. Patients in this group could be said to have had COPD ‘ruled out’ and therefore alternative explanations for their presenting symptoms could be sought. Since the N-Tidal^™^ device relies on 75 seconds of normal tidal breathing this categorization could be achieved at the point of care, in primary care.

Furthermore, the ability to rapidly identify those likely to have severe or very severe disease could ensure these patients receive prompt, intensive support (such as pulmonary rehabilitation or timely escalation of therapy as appropriate). The severity index offered by N-Tidal^™^ could also offer an alternative method to monitor disease progression. At a time when healthcare services even in high income countries can struggle to provide sufficient quality-assured spirometry to meet primary care needs, N-Tidal^™^ could alleviate the demand for further lung function testing for COPD for a significant portion of patients. In middle and low income settings where quality-assured spirometry is likely to remain out of reach of the majority of the population, N-Tidal^™^ could be a potential solution.

### Limitations and scope for future work

The analysis presented in this study has a number of limitations. First, simpler machine learning models were used in keeping with the National Health Service Artificial Intelligence recommendations regarding algorithmic explanability [15]. This limitation will likely be exacerbated as the size of high-resolution capnography datasets increase and opportunities to expore more complex fits to the data arise.

Second, ground-truth labels could only be obtained using current diagnostic pathways, known to have their shortcomings and inaccuracies. This is especially true for the less severe COPD patients who were included in this paper, whose rate of misdiagnosis is likely higher than GOLD 3 and 4 COPD patients. In addition, the GOLD severity labels used for this analysis were not provided by clinicians, but were inferred solely from percentage predicted FEV_1_ readings of patients with a diagnosis of COPD. In reality, other factors such as number of exacerbations per year, general respiratory symptoms and smoking history would likely also be taken into account when determining an overall impression of severity. For example, exacerbation frequency and symptom burden forms an integral part of GOLD’s ABE severity assessment tool. It is therefore possible that a misclassification or anomalous severity model output probability of a participant from one of the models presented in this article could be caused by mislabeling or misdiagnosis, particularly when considering the GOLD 1 COPD cohort.

The small sample size of the severity determination cohort and resultant removal of comorbid patients is another limitation and alongside the probably significant rate of GOLD 1 misclassification, decreases certainty on the model’s practicability. It will be possible to build on this work by collecting capnography data from more COPD patients, alongside severity labels which have been provided by clinicians based on spirometry that has been contextualized by the rest of the patient’s clinical history and CT scans. This will enhance label accuracy.

Finally, it is possible that the variety of disease data collected over the five studies and volunteer healthy cohort may fail to cover the full heterogeneity of lung conditions that would be encountered in a real-world clinical setting, limiting generalizability.

Regardless, the proposed methods managed to distinguish on capnography alone (without supplementary data such as smoking history), between participants with COPD and those with a range of plausible differential diagnoses for common symptoms of COPD (including healthy volunteers), demonstrating its potential clinical benefit.

In summary, we demonstrate that the N-Tidal^™^ fast-response capnometer and cloud analytics pipeline can perform real-time geometric waveform analysis and machine-learning-based classification to diagnose all severities of COPD. In contrast to commonly used ‘black box’ machine learning methodologies, a set of highly explainable methods were used that can provide traceability for machine diagnosis back to individual geometric features of the pCO2 waveform and their associated physiological properties suggestive of obstructive airways disease. In addition, the probability output of a separate machine learning model could be used as an alternative severity index for COPD using the N-Tidal^™^ device, with the interpretability of the implemented machine learning techniques providing a picture of COPD patients’ capnographic progression from GOLD 1 through GOLD 4.

## Data Availability

The datasets generated during and/or analyzed during the current study are not publicly available for data protection reasons.

## Acknowledgements

The ABRS study was supported by the National Institute for Health Research Invention for Innovation (NIHR i4i) Programme (Grant Reference Number: II-LA-1117-20002), the GBRS study was supported by Innovate UK (Grant Reference Number: 102977), the CBRS study was supported by SBRI Healthcare, and the CBRS2 study was supported by Pfizer OpenAir. The ABRS and GBRS research was supported by the Portsmouth Technology Trials Unit (www.pttu.org.uk) and the NIHR Community Healthcare MedTech and In Vitro Diagnostics Co-Operative (MIC). The CARES research was supported by the Modality GP Partnership (www.modalitypartnership.nhs.uk).The authors also acknowledge the work of staff at the Cambridge COPD Centre for Respiratory Research for their role in patient recruitment for CRBS and CBRS2, and staff in Oxford University’s Primary Care Clinical Trials Unit who were responsible for trial management (Julie Allen, Johanna Cook, Joy Rahman, Rebecca Edeson) and nursing (Heather Rutter, Karen Madronal, Bernadette Mundy) in ABRS. *BRS Study Team:* Jonathon Winter^2^, Andrew Gribbin^2^, Milan Chauhan^2^, Ruth De Vos^2^, Paul Kalra^2^, Selina Begum^2^, Barbara Robinson^3^, Bernadette Mundy^3^, Heather Rutter^3^ and Karen Madronal^3^.

## Author contributions

AC, TB, GH, DN, LW, and EV conceived of the individual studies and were responsible for study supervision and study design. DN, LW and EV collected the data. ABS, LT and AXP conceived of the original models, analyses and experimentation presented in this article. JCC and AXP designed and wrote the algorithms and code for denoising, pre-processing, and feature engineering. LT, ABS, HB, RHL, AXP wrote the code for, and carried out, model training and data analysis. AXP supervised the analyses in the manuscript. STW, AXP, GL, AC, TB, and GH contributed to expert technical review and data interpretation. LT, GL, ABS, HB, RHL drafted the manuscript. All authors participated in review and editing of the manuscript. All authors had access to the results data presented in the manuscript. The decision to submit this manuscript was made by all authors. All authors read and approved the final manuscript.

## Declarations

### Ethics approval and consent to participate

All participants consented to participate. Further details can be found on the National Library of Medicine’s Clinical Trial website for CBRS (NCT02814253), GBRS (NCT03356288), CBRS2 (NCT03615365), ABRS (NCT04504838) and CARES (NCT04939558).

### Consent for publication

Not applicable.

### Author details

^1^TidalSense Limited, 15a Vinery Rd, Cambridge CB1 3DN, UK. ^2^Portsmouth Hospitals University NHS Trust, Portsmouth, UK. ^3^Nuffield Department of Primary Care Health Sciences, NIHR Community Healthcare MedTech and IVD Cooperative, University of Oxford, Oxford, UK. ^4^Modality GP Partnership. ^5^Channing Division of Network Medicine, Department of Medicine, Harvard Medical School, Boston, MA, USA.

## References

[1] World Health Organisation. The top 10 causes of death, 2020.

[2] British Lung Foundation. Chronic obstructive pulmonary disease (copd) statistics, 2012.

[3] Amir Khakban, Don D. Sin, J. Mark FitzGerald, Bruce M. McManus, Raymond Ng, Zsuzsanna Hollander, and Mohsen Sadatsafavi. The projected epidemic of chronic obstructive pulmonary disease hospitalizations over the next 15 years. a population-based perspective. American Journal of Respiratory and Critical Care Medicine, 195(3):287–291, 2017. PMID: 27626508.

[4] NHS UK. Chronic obstructive pulmonary disease (copd) - treatment, 2017.

[5] Peter M A Calverley, Julie A Anderson, Bartolome Celli, Gary T Ferguson, Christine Jenkins, Paul W Jones, Julie C Yates, and Jørgen Vestbo. Salmeterol and fluticasone propionate and survival in chronic obstructive pulmonary disease, 2007.

[6] American Lung Association. Copd trends brief - burden, 2019.

[7] Amir Qaseem, Vincenza Snow, Paul Shekelle, Katherine Sherif, Timothy J. Wilt, Steven Weinberger, and Douglas K. Owens. Diagnosis and management of stable chronic obstructive pulmonary disease: a clinical practice guideline from the American College of Physicians. Annals of internal medicine, 147(9):633–638, nov 2007.

[8] M Bednarek, J Maciejewski, M Wozniak, P Kuca, and J Zielinski. Prevalence, severity and underdiagnosis of COPD in the primary care setting. Thorax, 63:402–407, 2008.

[9] Michael B. Jaffe. Using the features of the time and volumetric capnogram for classification and prediction. Journal of clinical monitoring and computing, 31(1):19–41, feb 2017.

[10] Tadukzwa Mkorombindo et al. Daiana Stolz. Towards the elimination of chronic obstructive pulmonary disease: a lancet commission. The Lancet Commissions, 400(10356):921–972, 2022. PMID: 36075255.

[11] Leeran Talker, Daniel Neville, Laura Wiffen, Ahmed B. Selim, Matthew Haines, Julian C. Carter, Henry Broomfield, Rui Hen Lim, Gabriel Lambert, Jonathon Winter, Andrew Gribbin, Milan Chauhan, Ruth De Vos, Paul Kalra, Selina Begum, Barbara Robinson, Bernadette Mundy, Heather Rutter, Karen Madronal, Scott T. Weiss, Gail Hayward, Thomas Brown, Anoop Chauhan, Ameera X. Patel, and BRS Study Team. Machine diagnosis of chronic obstructive pulmonary disease using a novel fast-response capnometer. Respiratory Research, 24(1):150, 2023.

[12] S.R. Bate, B. Jugg, S. Rutter, S. Graham, R. Perrott, R. Rendell, John Altrip, John Foord, Julian Carter, Matthew Haines, and J Walsh. N-Tidal C: A Portable, Hand Held Device for Assessing Respiratory Performance and Injury, 2018.

[13] Denis E. O’Donnell Peter Lange, David M Halpin and William MacNee. Diagnosis, assessment, and phenotyping of copd: beyond fev1. Int J Chron Obstruct Pulmon Dis., (11(Spec Iss)):3–12, 2016.

[14] Denis E. O’Donnell. Cardiovascular Institute of the South. Copd and heart failure: What are the symptoms and how are they related?, 2017.

[15] Indra Joshi and Jessica Morley. Artificial intelligence: How to get it right. putting policy into practice for safe data-driven innovation in health and care, 2019.

