## Supplementary Material for "Diagnosis and Severity Assessment of COPD using a Novel Fast-Response Capnometer And Interpretable Machine Learning"

[1] *TidalSense Limited, Cambridge, UK*
[2] *Portsmouth Hospitals University NHS Trust, Portsmouth, UK*
[3] *NIHR Community Healthcare MedTech and IVD Cooperative, Nuffield department of Primary Care Health Sciences, University of Oxford*
[4] *Modality GP Partnership*
[5] *Channing Division of Network Medicine, Department of Medicine, Harvard Medical School, MA, US*

---

### Feature Engineering

Reasons for capnogram exclusion included: nose breathing (incorrect breathing technique leading to low $CO_2$ values close to baseline), cardiogenic oscillations (high frequency fluctuations in $CO_2$ during the alveolar plateau occasionally observed during low frequency ventilation), condensation-compromised readings (indicated by abnormally high $CO_2$ values), breaths ended prematurely due to coughing or swallowing and overly noisy breaths. Viable breaths were moved on to feature extraction.

For each capnogram, 77 per-breath features and 5 whole capnogram features were derived. As there was a diverse number of breaths per capnogram and breath shape within a capnogram subtly varied, the median was calculated for the per-breath features in each capnogram. These features included the following: $\alpha$, $\beta$, $\gamma$, and $\delta$ angles (Figure 2, main text) [1, 2, 3]; gradients and residuals derived from fitting curves to phases, such as the expiratory plateau [4]; absolute and short-term variability of $pCO_2$ [5]; curvature and other higher-order time-based features such as the ratio of the expiratory to inspiratory phase [2, 3, 6]; second-order area ratios in quadrants of the expiratory phase and area under the curve (AUC), which are commonly calculated in volumetric capnography [6, 7]. Many of these features have been hypothesized to relate to clinical airway obstruction and have also been shown using machine learning to be effective at identifying salient physiological components of various obstructive airway diseases, including with the N-Tidal™ device [8].

Any breaths where the full feature set could not be calculated were also automatically excluded from analysis, and further checks were carried out manually to ensure that all condensation-compromised breaths had been excluded by automated methods.

### Machine Learning

Following pre-processing and waveform parameterization, each N-Tidal feature was normalized and scaled to a mean of zero and a standard deviation of 1. Significant class imbalances are known to produce misleadingly high overall accuracy while biasing machine learning models towards low predictive accuracy in the minority class [9]. To mitigate this risk, only six randomly- selected capnograms from each participant in the majority class (non-COPD) and fourteen randomly-selected capnograms from each participant in the minority class (COPD) were retained for the COPD classification task. This ensured the number of COPD and non-COPD capnograms in each class were equal.

The resultant machine learning capnogram dataset therefore represented only a fraction of the total breath records available. Given that data for some studies, such as CARES, was only collected for two weeks for each participant, the maximum number of capnograms used for training was limited to fourteen so that participants from studies with many

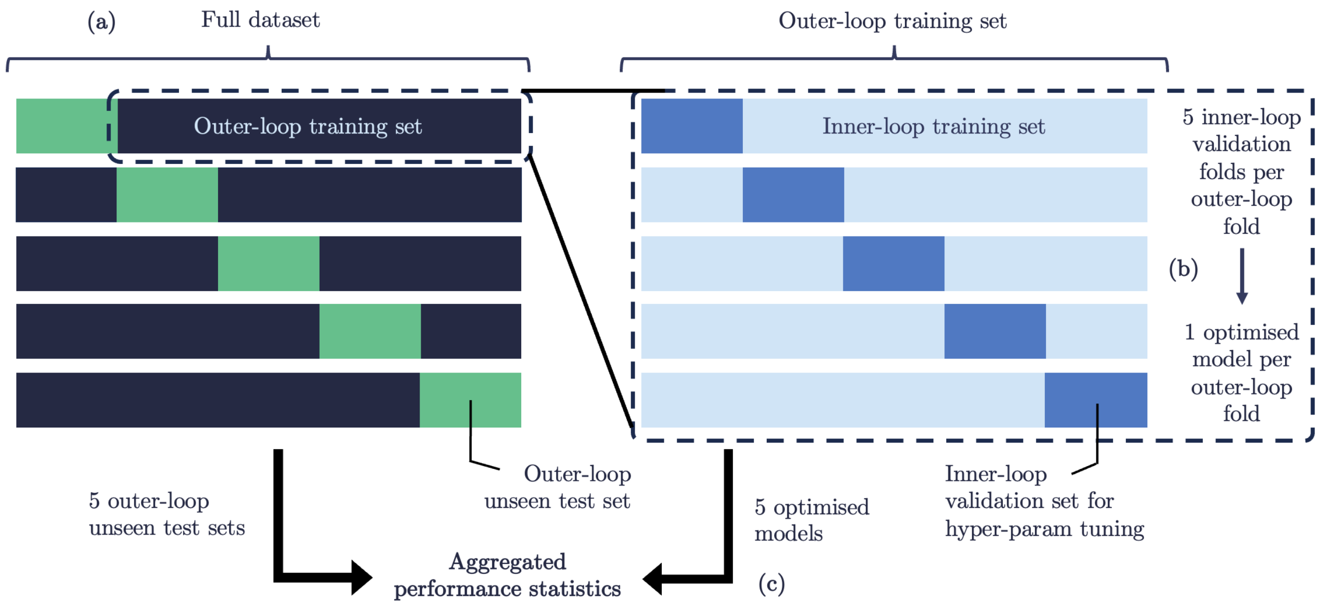


**Figure S1.** Overview of the nested cross-validation scheme used for training and model evaluation. (a) The full dataset is split into five outer-loop folds with non-overlapping patients. (b) For each outer-loop fold, an inner loop of five folds is run on the outer-loop training set to find optimal hyperparameters, and a model is subsequently trained on the full outer-loop training set. (c) The five optimized models from the five outer-loop training sets are tested on the five outer-loop test sets.

more capnograms per patient, such as CBRS, would not dominate the dataset. Limiting the number of capnograms per patient also ensured that the models would be less likely to overfit on features of individual patients such as confounding comorbidities, enabling them to learn the general capnographic characteristics of COPD across the whole population. Similarly, only fourteen randomly-selected capnograms from each participant in both classes (GOLD 1 and GOLD 4) were retained for the severity estimation task.

Next, model training and evaluation was undertaken using a nested cross-validation scheme (Figure S1). The full training dataset was first split into five 'outer-loop' folds using a group-stratified cross-validation procedure, ensuring the capnograms of a single patient would not be split across outer-loop training and test sets. For each outer-loop iteration, the outer-loop training set was re-split using group-stratified cross-validation into training and validation sets. An 'inner-loop' of five training/validation iterations was then run on the split outer-loop training set to find a set of hyperparameters which optimized performance. The full outer-loop training set was subsequently used to train a model using optimized hyperparameters, which was tested on the unseen outer-loop test set. This process was repeated once for each of five different outer-loop test sets and the results of the optimized models on all outer-loop test sets were aggregated to produce the results of the overall model performance. The performance variability across the five outer-loop test sets gave a measure of model generalizability.

The nested cross-validation scheme of training and evaluation had two distinct advantages. First, it maximized the data that was tested on as every patient was placed exactly once in an unseen outer-loop test set, which was especially important for the severity estimation task given the small number of patients in the dataset. Second, nested cross-validation mitigated the risk of optimistic model bias caused by tuning hyperparameters and testing the optimized model on the same test set.

For both the COPD diagnosis and severity determination tasks, the Python scikit-learn package was used to produce each of the following: sensitivity; specificity; negative predictive value (NPV); positive predictive value (PPV); micro-averaged area under ROC (AUROC); receiver operator characteristic (ROC) curves; and precision-recall (PR) curves. The most significant features driving model learning were extracted to understand which features of the capnogram waveform were most predictive of COPD and very severe COPD for the diagnostic and severity determination classifiers respectively.

### COPD Diagnosis

Demographic bias was also investigated as part of analysis of the COPD diagnostic model. First, misclassification rates were stratified by birth sex and COPD status (Table S1), highlighting that while there was a discrepancy in misclassification rates between men and women, these discrepancies were small (at most 6%, for the non-COPD cohort). Furthermore,

| | **Female** | **Male** | **Total** |
|---|---|---|---|
| **Non-COPD** | 16.4% (384/2343) | 22.0% (309/1403) | 17.3% (701/4053) |
| **COPD** | 22.9% (424/1851) | 18.6% (389/2100) | 20.7% (822/3965) |
| **Total** | 19.3% (808/4194) | 20.0% (698/3503) | 19.0% (1523/8018) |

**Table S1.** Misclassification rates of each sex versus disease group for the logistic regression (LR) model.

the difference between the medians of the age distributions of misclassified and correctly classified capnograms was not statistically significant (Mann-Whitney U=245, P = 0.23, two-tailed), indicating that age did not bias the rate of misclassification. Likewise, it was found that the difference between the medians of BMI distributions of misclassified and correctly classified examples were not statistically significant (Mann-Whitney U=230, P = 0.433, two-tailed) suggesting the same conclusion.

### Severity Determination

The regions of the capnogram that had the greatest ability to distinguish GOLD 1 and GOLD 4 COPD patients were investigated. The salient regions could also be interpreted as the regions which best describe the progression of COPD severity. The heatmap of waveform region importances (Figure S2) was obtained in the same manner as for the COPD diagnosis task. Features associated with the alpha (transition from expiratory upstroke to expiratory plateau) and beta (transition from expiratory plateau to inspiratory downstroke) regions of the waveform were found to be the most important drivers of learning. Significant importance was also ascribed to the phase 2 and phase 4a regions.

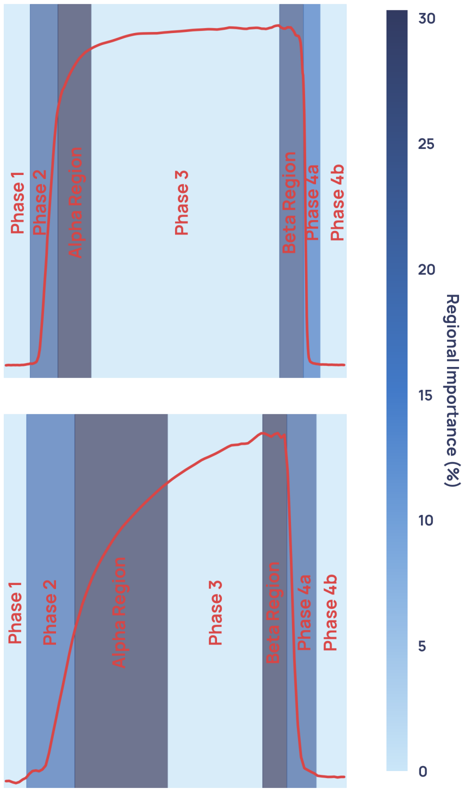


**Figure S2.** Average weighted feature importance by capnogram waveform region, where weighted features were calculated as the magnitude of the product of the normalized feature value and the feature importance. **(A)** shows an example for a GOLD 1 waveform, and **(B)** shows an example for a GOLD 4 waveform.

### Model Robustness

For the diagnostic task, the best performing model over the five unseen outer-loop test sets was support vector machine (SVM), with a class-balanced AUROC of 0.811 $\pm$ 0·022 and positive predictive value (PPV) of 0·834 $\pm$ 0·015. The robustness of the COPD diagnostic models was demonstrated in three ways.

First, there was only a 0.05 difference in accuracy between the best-performing (SVM with RBF kernel) and worst-performing (XGBoost) models. This affirms that the accurate classification of COPD and non-COPD patients was rooted in a strong underlying signal in the data itself. The linear algorithm (LR) performed almost as well as SVM and outperformed XGBoost, suggesting that simple fits to the feature set capture as much information that can be used to distinguish the classes as more complex fits do.

Second, the mean training accuracies over the five outer-loop training sets for LR, XGBoost and SVM were 0.819, 0.984 and 0.858. The relatively small differences between train and test performance for the LR (simplest, most explainable and second highest-performing) and SVM (highest-performing) models in particular demonstrate their generalizability and indicates that the models were not overfitting to the training or test sets during the five-fold outer-loop cross-validation, as the outer-loop test sets used data from completely unseen patients. This gives confidence in the model's efficacy in a real diagnostic scenario.

Finally, the variability of unseen test set performance between the outer-loop folds gives an indication of the models' abilities to generalize to different combinations of training and test data. The highest standard deviation that was seen in any performance metric across the folds was 0.055, whilst the majority of metrics (including diagnostic accuracy for all models) had standard deviation in test performance of less than 0.03. This reinforces the applicability of the model to unseen testing data, as well as its ability to maintain strong test performance through periodic retraining after deployment in clinical practice.

The severity determination LR model demonstrated similar robustness to the COPD diagnostic models. Mean test and train performance across the five outer-loop folds were both very strong at 0.959 and 0.979 respectively, with the relatively small difference between them demonstrating the model did not overfit training data. Standard deviation of model performance over the folds in some metrics was higher those seen in the COPD diagnostic task, but this is to be expected given the much smaller number of patients in the dataset (43 patients for severity, 999 patients for COPD diagnosis), meaning that each group-stratified train/test set was a less accurate approximation of the overall dataset. Despite the small number of patients, standard deviations across folds in accuracy (0.039), AUROC (0.013) and PPV (0.039) were all indicative of generalizability.

### Further Study Details

#### COPD Breathing Record Study (CBRS)

##### Primary Objective

To collect a longitudinal observation study database of capnograph records for up to 30 patients with COPD over 6 weeks using the N-Tidal C data-collector capnometer.

##### Secondary Objectives

- To identify the correlation of carbon dioxide ($CO_2$) measurements by capnography with those obtained by periodic standard arterial and capillary blood gas measurements.
- To identify the within-day and day-to-day variability of exhaled carbon dioxide in patients with COPD.
- To measure the absolute change in exhaled carbon dioxide measurements of patients admitted with an acute exacerbation of COPD during their recovery.
- To capture the carbon dioxide signature predictive of exacerbation in monitored patients who undergo an acute admission during the study.

##### Device Specific Objectives

- To assess the ability of patients with COPD to use the N-Tidal C data-collector capnometer daily and to capture user feedback.
- To assess the frequency of replacement of the consumable breath tubes and mouthpieces in normal daily use.

**Inclusion criteria**

- Aged 18 years and over.
- Case managed group only: Diagnosis of COPD; Chronically elevated $PaCO_2$; Susceptible to frequent exacerbations of COPD
- Acute admission group only: Hospitalized via the emergency room for treatment of COPD-related ventilatory failure.
- Provided written, informed consent.

**Exclusion criteria**

- Diagnosis of neuromuscular disorders or kyphoscoliosis.
- Patients who, in the opinion of the investigator, are unlikely to comply with the requirements of the study, use the device correctly, or keep the diary records.

#### General Breathing Record Study (GBRS)

**Primary Objective**

To explore the characteristics of the Tidal Breathing carbon dioxide ($TBCO_2$) waveform that can differentiate between different respiratory and cardiac conditions (including acute and chronic disease states) and establish a profile for healthy controls.

**Secondary Objectives**

- To identify within-patient changes in the $TBCO_2$ waveform that may predict or detect a deterioration of the underlying disease.
- To establish whether the characteristics of any waveform changes before a deterioration are similar in all patients in the same group.
- To identify waveform features that may help inform a larger disease-specific prospective study.
- To describe the relationship between characteristics of the $TBCO_2$ waveform and severity of the primary condition of interest (as measured by disease-specific clinical parameters and symptom questionnaires).
- To compare the use of the $TBCO_2$ waveform (and the N-Tidal C device) in monitoring different breathing conditions to traditional methods of monitoring disease control.
- To identify the correlation of carbon dioxide ($CO_2$) measurements by capnography with those obtained by periodic standard arterial blood gas measurements in patients with respiratory diagnoses.
- To monitor the safety of the device in regular use by patients at home.
- To establish the 'ease of use' of capnography measurement as a potential disease diagnosis and monitoring method in all participants.
- To establish the 'ease of use' of capnography measurement in HCP's.
- To identify the ranges of $TBCO_2$ waveform parameter values (minimum and maximum) for the different disease cohorts.
- To identify within-day and day-to-day variations in $TBCO_2$ waveform parameter values for the different disease cohorts.

**General Inclusion Criteria**

- Male or female, aged $\geq$ 16 years
- Willing and able to provide written informed consent.

**General Exclusion Criteria**

- Known other lung, chest wall, neuromuscular, cardiac or other comorbidity or abnormality that would affect spirometry and/or other measures of lung function or $TBCO_2$ measurements.

- In the opinion of the clinical investigator, the participant would have difficulty completing the study procedures consistently over 6 months.

**Asthma Inclusion Criteria**

- A confirmed clinical diagnosis of asthma for $\geq$ 6 months supported by evidence of any of the following:
  - Airflow variability, with a variability in FEV1 of $>$20% across clinic visits within the preceding 12 months, with concomitant evidence of airflow obstruction (FEV1/FVC ratio $<$70% on spirometry).
  - Airway reversibility with an improvement in FEV1 by $\geq$ 12% or 200 ml after inhalation of 400 $\mu$g of salbutamol via a metered dose inhaler and spacer within the preceding 12 months.
  - Airway hyper-responsiveness demonstrated by Methacholine (or similar) challenge testing with a provocative concentration of Methacholine required to cause a 20% reduction in FEV1 (PC20) of $\leq$ 8mg/ml or equivalent test.

- Moderate to severe asthma defined as BTS stage 3 5

- Exacerbation free for $>$2 weeks (defined as no increased dose or course of oral corticosteroids or antibiotics).

- 2 or more exacerbations in the previous 12 months with at least 1 exacerbation within the last 6 months.

**Breathing Pattern Disorder / Vocal Cord Dysfunction Inclusion Criteria**

A Clinical diagnosis of a Breathing Pattern Disorder (BPD) by a Specialist Respiratory Physiotherapist.

**Chronic Heart Failure**

- A confirmed clinical diagnosis of chronic heart failure with both of the following:
  - A Left Ventricular Ejection Fraction $<$40% on most recent imaging within the last 12 months.
  - New York Heart Association Class 2-4

- Either (i) admitted with an acute decompensation of their heart failure to hospital requiring intravenous diuretics or an increase in diuretic dose from baseline (e.g., 40mg or more furosemide) within the last 6 months or (ii) stable outpatient with NT-proBNP $>$400ng/mL in sinus rhythm or NT-proBNP $>$1000ng/mL in atrial fibrillation.

**Motor Neurone Disease**

A confirmed clinical diagnosis of Motor Neuron Disease (MND)

**Pneumonia**

A confirmed clinical diagnosis of Pneumonia supported by evidence of consolidation on a chest X-ray (CXR) or computed tomography (CT) imaging.

**Healthy Volunteers**

- No known history of lung, cardiac or neuromuscular disease (defined as no current clinical diagnosis of, or receiving treatment for, a lung, cardiac or neuromuscular disease).

- BMI $\leq$ 40

- Non-smoker, or ex-smoker with $\leq$ 5 pack year history

### COPD Breathing Record Study 2 (CBRS2)

**Primary Objective**

To assess the changes in key parameters in the tidal breathing $CO_2$ ($TBCO_2$) waveform, including the $\alpha$ angle, the minimum $CO_2$ level achieved and the stability of the expiratory cycle, during the transition from stable COPD to during acute exacerbations.

**Secondary Objectives**

- To collect a longitudinal observational database of $TBCO_2$ waveform records for up to 50 patients with moderate-to-severe COPD over 26 weeks using the N-Tidal C Data Collector Device.
- To capture the carbon dioxide signature predictive of exacerbations in monitored patients who undergo mild, moderate and severe exacerbations during the study.
- To identify the within-day and day-to-day variability of respired $CO_2$ in patients with COPD.

**Inclusion Criteria**

- Aged 40 years and over.
- Diagnosis of COPD (Primary) and at least one moderate exacerbation within 12 months of starting the study period.
- Able to provide signed informed consent.

**Exclusion Criteria**

- Patients who, in the opinion of the investigator, are unlikely to comply with the requirements of the study, use the device correctly or keep the diary records.
- Diagnosis of neuromuscular disorders or Kyphoscoliosis.
- Diagnosis of other Respiratory disorders that, in the investigator's opinion, would impact the conduct of the study e.g., clinically significant bronchiectasis, asthma.
- Patients who have experienced an exacerbation of their COPD that has required treatment with antibiotics and/or oral corticosteroids within 2 weeks before the study start.

#### Asthma Breathing Record Study (ABRS)

**Primary Objective**

To determine characteristics within the $TBCO_2$ waveform shape, as measured by the N-Tidal C data collector device, that identify deteriorations in the user's respiratory condition, including changes leading to asthma exacerbations, and discriminate between poorly and well-controlled asthma.

**Secondary Objectives**

- To determine whether changes in the $TBCO_2$ waveform, measured by the N-Tidal C data collector device, can predict asthma exacerbations.
- To describe the relationship between characteristics of the $TBCO_2$ waveform and severity of asthma at baseline, as measured by:
  - BTS Stage 2-5
  - Disease control (Asthma Control Questionnaire)
  - Quality of Life (Asthma Quality of Life Questionnaire)
  - Spirometry (% predicted FEV1)
  - Fractional exhaled Nitric Oxide (FeNO in ppb, if available)
  - Airway resistance (Airway Oscillometry, if available)
  - Peak Expiratory Flow (PEF)
  - Forced Expiratory Flow (FEF25-75)
- To analyze subgroups of exacerbations according to the presence of triggers.
- To explore the feasibility of monitoring asthma control at home using the N-Tidal C in all participants.
- To assess the usability and acceptability of the device to study participants and their family/carers, gathering ideas for improved use and further development.
- To assess the adherence to use of the device by participants and explore barriers and facilitators to adherence.
- To evaluate healthcare resource use, costs and quality-of-life measures over the study period.
- To describe and quantify any adverse device effects.

**Inclusion Criteria**

- Male or Female, aged $\geq$ 7 years.
- Confirmed clinician diagnosis of asthma by examination of medical records and based on accepted national and/or international criteria e.g., BTS/SIGN, or GINA
- Moderate or Severe asthma (defined as BTS stage 2-5)
- Poorly controlled asthma (defined as an ACQ score of $\geq$ 1)
- Exacerbation prone asthma (defined as at least 1 asthma exacerbation requiring oral corticosteroid treatment in the last 12 months)
- Capable of providing written informed consent, or parental/guardian consent and participant assent in the case of a child

**Exclusion Criteria**

- Inability to understand or comply with study procedures and/or give fully informed consent.
- Known other lung, chest wall, neuromuscular, cardiac or other comorbidity or abnormality that would affect spirometry and/or other measures of lung function or $TBCO_2$ measurements (including Breathing Pattern Disorder or Chronic Obstructive Pulmonary Disease).
- Smokers (current or ex-smokers) with a >10 pack year history.
- In the opinion of the clinical investigator, the participant would have difficulty completing the study procedures consistently (for example, difficulty holding the device, or long periods of absence/travel) throughout the study period.

#### Cardiorespiratory Diagnostic Study (CARES)

**Primary objective**

This study uses a new breathing device called 'N-Tidal C' handset which measures breathing patterns. Investigators have found that people with cardiac and respiratory illnesses breathe out a gas, called carbon dioxide ($CO_2$), in a different way to healthy people. The pattern of breathed out $CO_2$ (the waveform) varies according to the underlying health of the user's lungs. Monitoring these changes may help doctors to more accurately diagnose and monitor the most common and serious respiratory conditions.

**Inclusion Criteria**

- Age >18 years
- Healthy volunteer (with no previous or current chronic cardiorespiratory diagnoses)

One of the following cardiorespiratory diagnoses:

- COPD (GOLD 1, 2, 3 / A, B, C)*
- Asthma (mild to moderate, not labeled as severe)*
- Congestive cardiac failure*
- Anaemia (with at least 50% of participants recruited having no history of chronic cardiorespiratory conditions)*
- Bronchiectasis (acquired or genetic, e.g. cystic fibrosis or other primary ciliary dyskinesias)*
- Lung cancer (including rare types e.g. mesothelioma)*
- Interstitial Lung Disease (including pulmonary fibrosis pneumoconiosis, asbestosis, sarcoidosis, amyloidosis)*
- Long COVID*
- Upper airway obstruction disorder*
- (Active pulmonary hypertension)
- (Extrinsic Allergic Alveolitis)
- (Active pulmonary embolism)

**Exclusion Criteria**

- Participants who, in the opinion of the chief investigator, or their delegate, are unlikely to comply with the requirements of the study.
- Diagnosis of neuromuscular disorders.
- Concurrent diagnosis of cardiorespiratory conditions (other than those listed in the inclusion criteria above) that, in the opinion of the chief investigator, would impact the conduct of the study.
- Participants who are acutely unwell, e.g. active exacerbation, very short of breath e.g. severe COPD (GOLD 4 / D) or end-stage IPF.
- Inability to give written informed consent.

| | COPD Breathing Record Study (CBRS) | General Breathing Record Study (GBRS) | COPD Breathing Record Study 2 (CBRS2) | Asthma Breathing Record Study (ABRS) | Cardiorespiratory Diagnostic Study (CARES) |
|---|---|---|---|---|---|
| **Primary Objective** | To collect a longitudinal observation study database of capnograph records for up to 30 patients with recent or recurrent exacerbations of COPD over 6 weeks using. | To explore the characteristics of the $TBCO_2$ waveform that can differentiate between different respiratory and cardiac conditions (including acute and chronic disease states) and establish a profile for healthy controls. | To assess the changes in key parameters in the $TBCO_2$ waveform, including the $\alpha$ angle, the minimum $CO_2$ level achieved and the stability of the expiratory cycle, during the transition from stable COPD to during acute exacerbations. | To determine characteristics within the $TBCO_2$ waveform shape that identify deteriorations in the user's respiratory condition, including changes leading to asthma exacerbations, and discriminate between poorly and well-controlled asthma. | To demonstrate that characteristics of the TBCO2 waveform, as measured by the N-Tidal C device, can be used to accurately classify individuals with COPD (including mild-to-moderate COPD) as distinct from those without COPD (i.e. those with other cardiorespiratory conditions or no history of cardiorespiratory conditions). |
| **Population** | Community group - chronically elevated $CO_2$ and frequent exacerbations of COPD; Acute admissions group – admitted to hospital with an exacerbation of COPD. | Disease cohorts selected from hospital outpatient clinic lists and inpatient wards. Healthy volunteers. | Patients with moderate-to-severe COPD from the Cambridge COPD center. | Patients with poorly controlled asthma. | Patients from the primary care patient population served by the Modality (GP) partnership. |
| **Clinical condition(s)** | COPD | Asthma, breathing pattern disorder, chronic heart failure, motor neuron disease, pneumonia, healthy volunteers. | COPD | Asthma | COPD (GOLD 1, 2, 3), Asthma (mild to moderate, not labeled as severe), Congestive cardiac failure, Anaemia, Bronchiectasis, Lung cancer, Interstitial Lung Disease, Long COVID, Upper airway obstruction disorder, Active pulmonary hypertension, Active pulmonary embolism, Extrinsic Allergic Alveolitis. |
| **Number of participants** | COPD: 30 | Asthma: 20; BPD, CHF, MND, Pneumonia, Healthy 10 each; Total: 70 | COPD: 50 | Asthma: 124; 92 of these were recruited from primary care, and 32 from secondary care. | Anemia: 95; Asthma: 134; Bronchiectasis: 30; COPD: 236; Healthy: 43; Heart Failure: 64; Long COVID: 53; Lung Cancer: 7; Other: 2; Pulmonary Fibrosis: 17; Upper Airway Obstruction: 55 |
| **Location** | Addenbrooke's Hospital, Cambridge University Hospitals NHS Foundation Trust, UK | Queen Alexandra Hospital and specialist secondary care community clinics, Portsmouth Hospitals University NHS Trust, UK | COPD Centre, Addenbrooke's Hospital, Cambridge University Hospitals NHS Foundation Trust, UK | Queen Alexandra Hospital, Portsmouth Hospitals University NHS Trust, UK and GP practices, Oxford, UK | 56 GPs across England |
| **Recruitment setting** | Outpatient, Inpatient | Outpatient | Outpatient | Outpatient, Inpatient and Primary Care | Primary care |
| **Duration** | 17th Feb 2016 – Dec 2016 | 9th Aug 2017 – 4th Jul 2018 | 15th Aug 2017 – 23rd Nov 2018 | 11th Feb 2020 – 31st Jan 2022 | 8th June 2021 - 1st November 2022 |
| **Clinical Trials.gov Identifier** | NCT02814253 | NCT03356288 | NCT03615365 | NCT04504838 | NCT04939558 |
| **Number of capnograms** | 2620 | 15803 | 14885 | 38026 | 25118 |
| **Additional data collected** | Medical history, clinical assessment, demographics, vital signs, spirometry, routine blood tests, blood gases. | Medical history, clinical assessment, demographics, vital signs, spirometry (not heart failure or pneumonia groups). Other assessments were disease specific. | Medical history, clinical assessment, demographics, symptoms, vital signs, spirometry. | Medical history, clinical assessment, demographics, vital signs, spirometry (plus optional others - FeNO, oscillometry, full body plethysmography) routine blood tests. | Medical history, clinical assessment, demographics, vital signs, spirometry, routine blood tests, blood gases. |

**Table S2.** Summary of the five clinical studies from which the paper has drawn its data.

### References

[1] C. L. Herry, D. Townsend, G. C. Green, A. Bravi, and A. J.E. Seely. Segmentation and classification of capnograms: application in respiratory variability analysis. *Physiological Measurement*, 35(12):2343, nov 2014.

[2] Tan Teik Kean, A. H. Teo, and M. B. Malarvili. Feature extraction of capnogram for asthmatic patient. *2010 2nd International Conference on Computer Engineering and Applications, ICCEA 2010*, 2:251–255, 2010.

[3] Barak Pertzov, Michal Ronen, Dror Rosengarten, Dorit Shitenberg, Moshe Heching, Yael Shostak, and Mordechai R. Kramer. Use of capnography for prediction of obstruction severity in non-intubated COPD and asthma patients. *Respiratory Research*, 22(1):1–9, dec 2021.

[4] Abubakar Abid, Rebecca J Mieloszyk, George C Verghese, Baruch S Krauss, and Thomas Heldt. Model-Based Estimation of Respiratory Parameters from Capnography, with Application to Diagnosing Obstructive Lung Disease. *IEEE Transactions on Biomedical Engineering*, 64(12):2957–2967, dec 2017.

[5] Huib R. Van Genderingen, Nikolaus Gravenstein, Jan J. van der Aa, and Joachim S. Gravenstein. Computer-assisted capnogram analysis. *Journal of Clinical Monitoring 1987 3:3*, 3(3):194–200, jul 1987.

[6] Karl Z. Lukic, Bruce Urch, Michael Fila, Marie E. Faughnan, and Frances Silverman. A novel application of capnography during controlled human exposure to air pollution. *BioMedical Engineering Online*, 5(1):1–11, oct 2006.

[7] J. A. Kline and M. Arunachlam. Preliminary study of the capnogram waveform area to screen for pulmonary embolism. *Annals of Emergency Medicine*, 32(3 I):289–296, 1998.

[8] Leeran Talker, Daniel Neville, Laura Wiffen, Ahmed B. Selim, Matthew Haines, Julian C. Carter, Henry Broomfield, Rui Hen Lim, Gabriel Lambert, Jonathon Winter, Andrew Gribbin, Milan Chauhan, Ruth De Vos, Paul Kalra, Selina Begum, Barbara Robinson, Bernadette Mundy, Heather Rutter, Karen Madronal, Scott T. Weiss, Gail Hayward, Thomas Brown, Anoop Chauhan, Ameera X. Patel, and BRS Study Team. Machine diagnosis of chronic obstructive pulmonary disease using a novel fast-response capnometer. *Respiratory Research*, 24(1):150, 2023.

[9] Shaza M Abd Elrahman and Ajith Abraham. A Review of Class Imbalance Problem. *Journal of Network and Innovative Computing*, 1:332–340, 2013.
